# Comparing Machine Learning Algorithms for Predicting ICU Admission and Mortality in COVID-19

**DOI:** 10.1101/2020.11.20.20235598

**Authors:** Sonu Subudhi, Ashish Verma, Ankit B. Patel, C. Corey Hardin, Melin J. Khandekar, Hang Lee, Triantafyllos Stylianopoulos, Lance L. Munn, Sayon Dutta, Rakesh K. Jain

## Abstract

As predicting the trajectory of COVID-19 disease is challenging, machine learning models could assist physicians determine high-risk individuals. This study compares the performance of 18 machine learning algorithms for predicting ICU admission and mortality among COVID-19 patients. Using COVID-19 patient data from the Mass General Brigham (MGB) healthcare database, we developed and internally validated models using patients presenting to Emergency Department (ED) between March-April 2020 (n = 1144) and externally validated them using those individuals who encountered ED between May-August 2020 (n = 334). We show that ensemble-based models perform better than other model types at predicting both 5-day ICU admission and 28-day mortality from COVID-19. CRP, LDH, and procalcitonin levels were important for ICU admission models whereas eGFR <60 ml/min/1.73m^2^, ventilator use, and potassium levels were the most important variables for predicting mortality. Implementing such models would help in clinical decision-making for future COVID-19 and other infectious disease outbreaks.

## Introduction

The COVID-19 pandemic has led to significant morbidity and mortality throughout the world ^1^. The rapid spread of SARS-CoV-2 has provided limited time to identify factors involved in SARS-CoV-2 transmission, predictors of COVID-19 severity, and effective treatments. At the height of the pandemic, areas with high number of SARS-CoV-2 infections were resource-limited and forced to ration life-saving therapies such as ventilators and dialysis machines ^2,3^. In this setting, being able to identify patients requiring intensive care or at high risk of mortality upon presentation to the hospital may help providers expedite patients to the most appropriate care setting.

Model predictions are gaining increasing interest in clinical medicine. Machine learning applications have been used to help predict acute kidney injury ^4^ and septic shock ^5^, amongst other outcomes in hospitalized patients. These tools have also been applied to outpatients to predict outcomes such as heart failure progression ^6^. Machine learning tools can be applied to predict outcomes such as Intensive Care Unit (ICU) admission and mortality ^7^. Thus far there have been few studies that examined specific machine learning algorithms in predicting outcomes such as mortality in COVID-19 patients ^8-10^. Given the potential utility of machine learning-based decision rules and the urgency of the pandemic, a concerted effort is being made to identify which machine learning applications are optimal for given sets of data and diseases ^11^.

To address this knowledge gap, we conducted a multi-hospital cohort (Mass General Brigham (MGB) healthcare database) study to extensively evaluate the performance of 18 different machine learning algorithms in predicting ICU admission and mortality. Our goal was to identify the best prognostication algorithm using demographic data, comorbidities, and laboratory findings of COVID-19 patients who visited emergency departments (ED) at MGB between March and April 2020. We validated our models on a temporally distinct patient cohort that tested positive for COVID-19 and had ED encounter between May and August 2020. We also identified critical variables utilized by the model to predict ICU admission and mortality.

## Results

### Patient characteristics

We obtained data from 10,826 patients in the multihospital database (Massachusetts General Brigham Healthcare database) who had COVID-19 infection during the period of March and April 2020. A total of 3,713 out of the 10,826 patients visited EDs. We evaluated patients based on demographics, medication use, history of past illness, clinical features, and laboratory values described in Table S1. After excluding patients with missing data, 1,144 patients remained, 99% of which were in-patients (n = 1133). For external validation, we pulled data of temporally distinct individuals from the Mass General Brigham (MGB) healthcare database who were positive for SARS-CoV-2 between May and August 2020. During this period, 1,754 out of 8,013 SARS-CoV-2 positive individuals visited the ER. After excluding patients with missing variables from Table S1, a total of 334 patients were left (98% of which were in-patients).

The baseline characteristics of 1,144 patients in the training dataset are listed in Table 1. The overall study population included 45% women, and the majority were above the age of 60. The number of patients who were admitted to ICU within 5 days and who died within 28 days of ED visit were 342 (30%) and 217 (19%), respectively. The external validation dataset included patients with similar distribution in age ≥50 years (X^2^(4, N = 1193) = 8.9, p = 0.063), gender (X^2^ _(1, N = 1478)_ = 0.017, p = 0.89), race (X^2^ _(1, N = 1478)_ = 0.07, p = 0.79) and BMI (X^2^ _(2, N = 1478)_= 4.31, p = 0.12) (Table S6). Of the 334 patients who visited the ED, 74 (22%) were admitted to the ICU and 45 (13%) died with COVID-19.

**Table 1.**
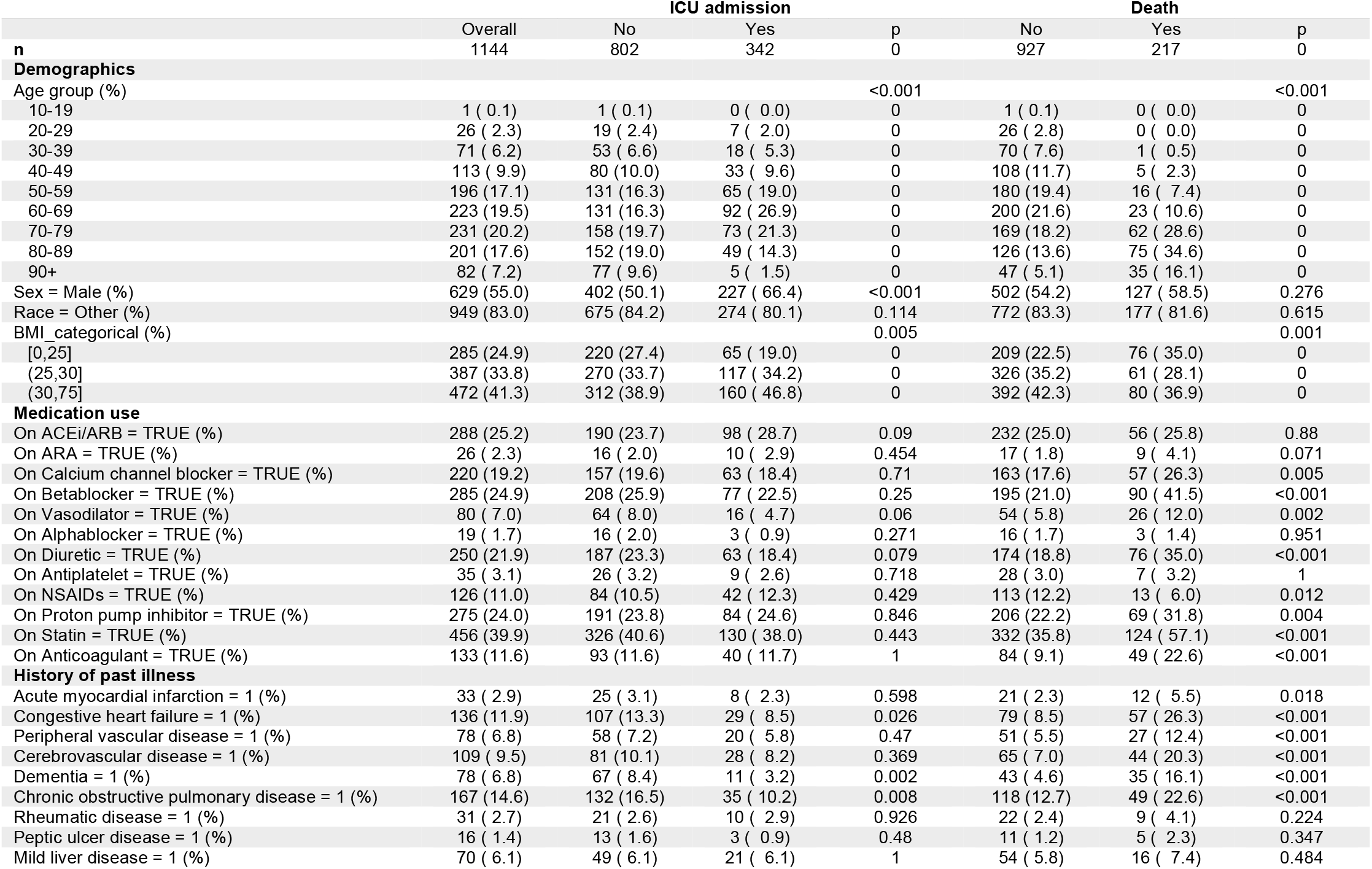
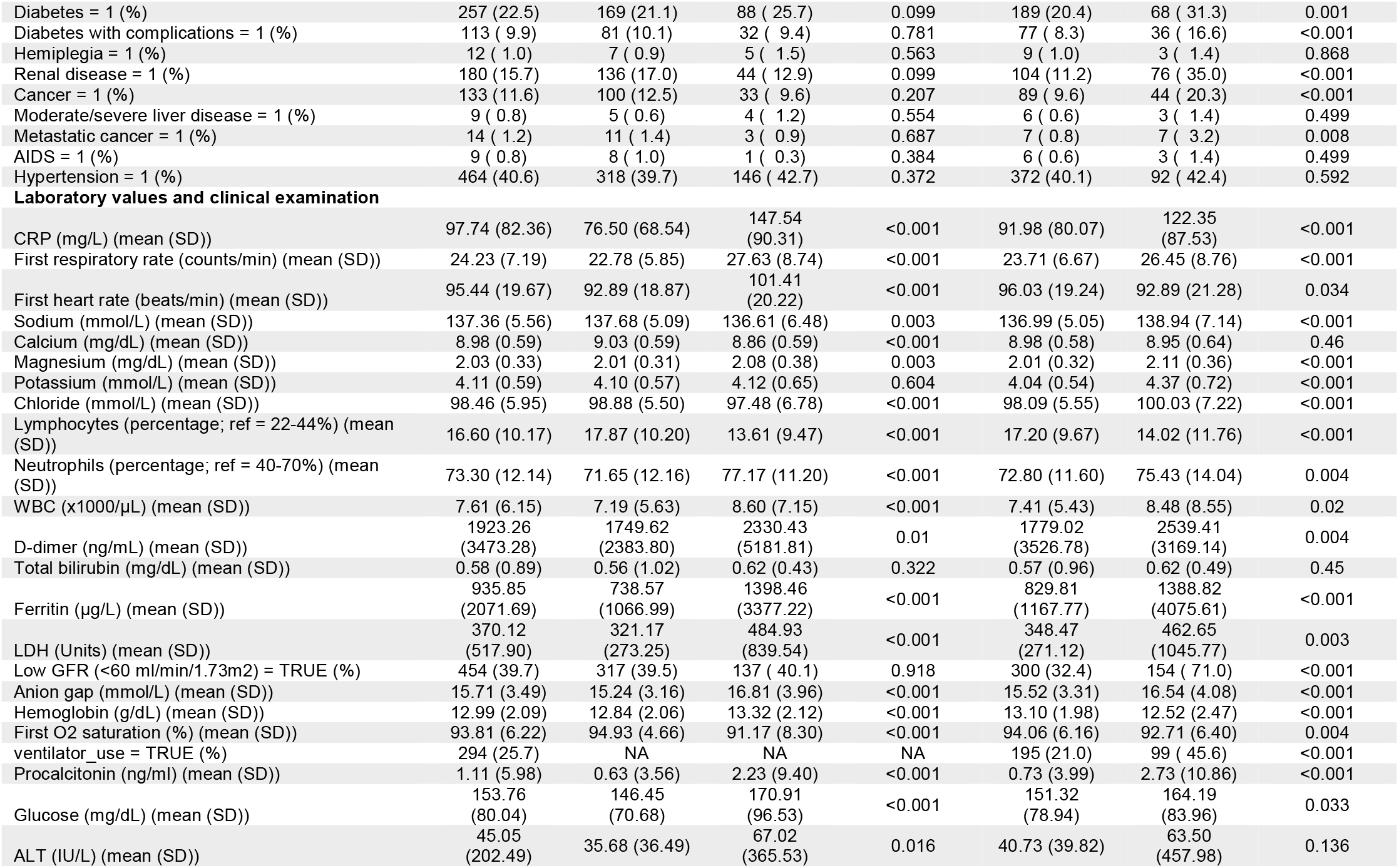
Characteristics of patients who visited emergency department during March and April 2020 for COVID-19, that were included for building the machine learning models. Variables stratified based on ICU admission and death of patients.

### Comparing performance of prediction models – cross validation

We evaluated 18 machine learning algorithms belonging to 9 broad categories, namely ensemble, Gaussian process, linear, naïve bayes, nearest neighbor, support vector machine, tree-based, discriminant analysis and neural network models.

On comparing the ICU admission prediction models using cross validation, we observed that all ensemble-based models had mean precision-recall area under curve (PR AUC) scores more than 0.77 (Table 2; Fig. S2A-B). Specifically, the PR AUC score for *AdaBoostClassifier* was 0.80 (95% CI, 0.73 – 0.87), for *BaggingClassifier* was 0.80 (95% CI, 0.73 – 0.87), for *GradientBoostingClassifier* was 0.77 (95% CI, 0.68 – 0.86), for *RandomForestClassifier* was 0.80 (95% CI, 0.70 – 0.90), for *XGBClassifier* was 0.78 (95% CI, 0.70 – 0.86), and for *ExtraTreesClassifier* was [0.79 (95% CI, 0.72 – 0.86)]. In addition, *LogisticRegression* [0.79 (95% CI, 0.71 – 0.87)], and *LinearDiscriminantAnalysis* [0.76 (95% CI, 0.68 – 0.84)] also had high PR AUC scores. In contrast, *GaussianProcessClassifier* [0.6 (95% CI, 0.54 – 0.66)], *SGDClassifier* [0.63 (95% CI, 0.60 – 0.66)] and LinearSVC [0.65 (95% CI, 0.57 – 0.73)] had low PR AUC scores. Upon performing multiple comparison analysis between all models (based on PR AUC and ROC AUC scores), the ensemble-based models and *LogisticRegression* models have similar pattern of performance (Fig. S1A-B). On grouping the models based on their broad categories, we found that ensemble models have significantly higher PR AUC scores than all other model types except for logistic regression (based on Fisher’s Least Significant Difference (LSD) t-test; Fig. 2A; details of statistical analysis in Table S7). For ROC AUC scores, ensemble models performed better than all models except logistic regression (Fig. 2A; Table S7).

**Table 2.**
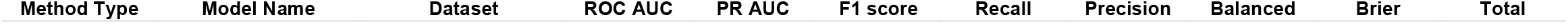
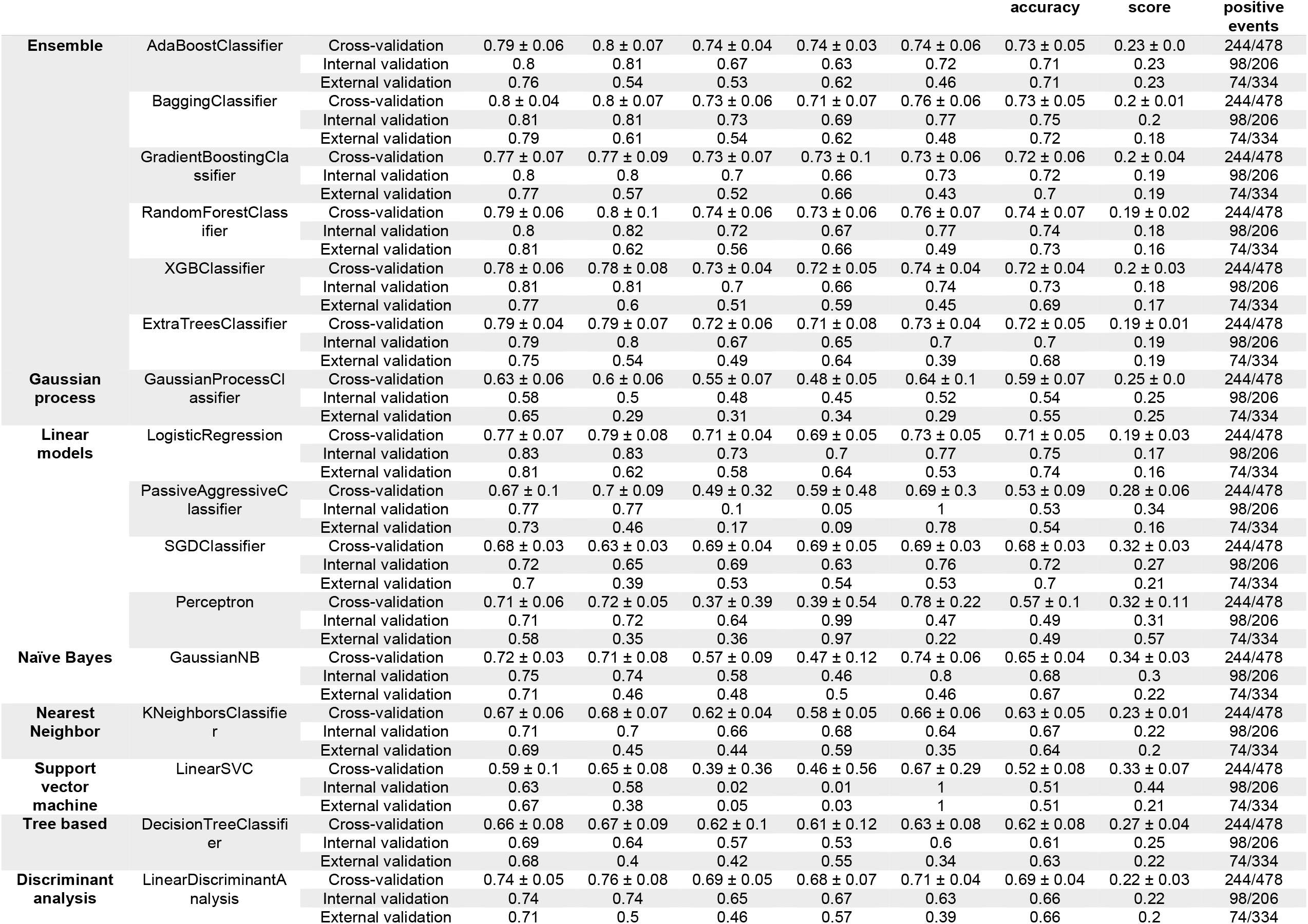
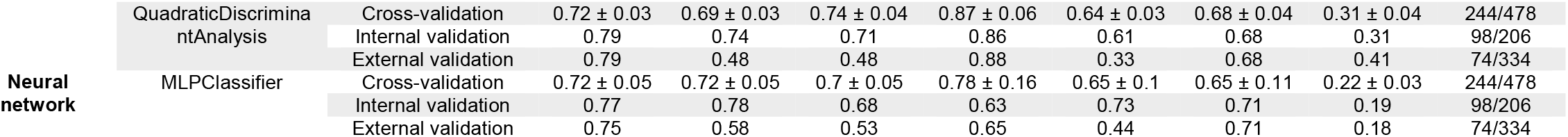
Performance of machine learning models to predict ICU admission within 5 days in COVID-19 patients. Cross-validation scores are expressed as mean ± 95% confidence interval.

**Fig. 1.**
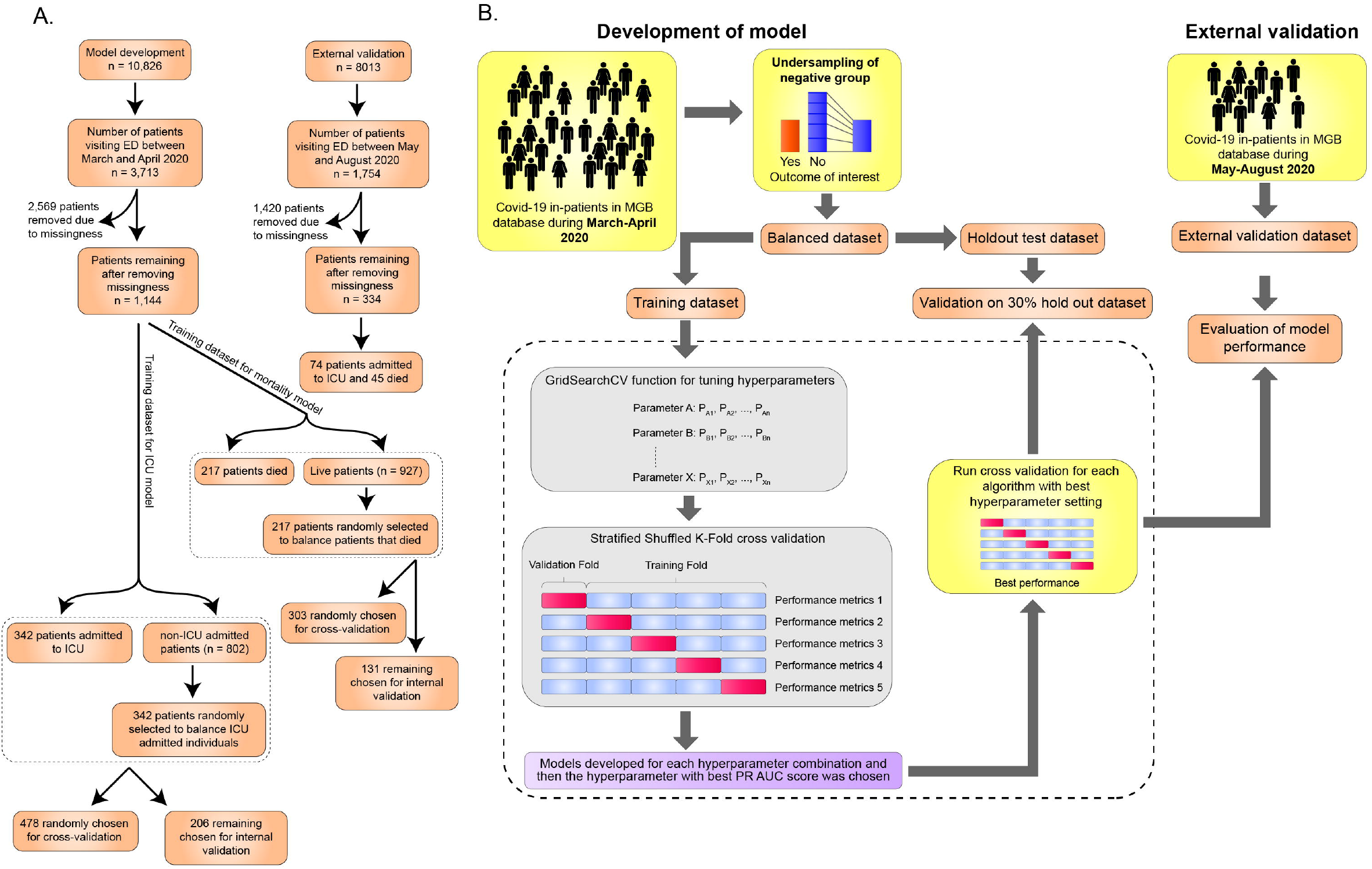
Schematic diagram representing the process of machine learning model development. **(A)** Flow diagram depicting steps in obtaining the training and external validation datasets (with patient numbers in each step). **(B)** The process of patient selection, dataset balancing, hyperparameter tuning, cross-validation, internal and external validation are shown.

**Fig. 2.**
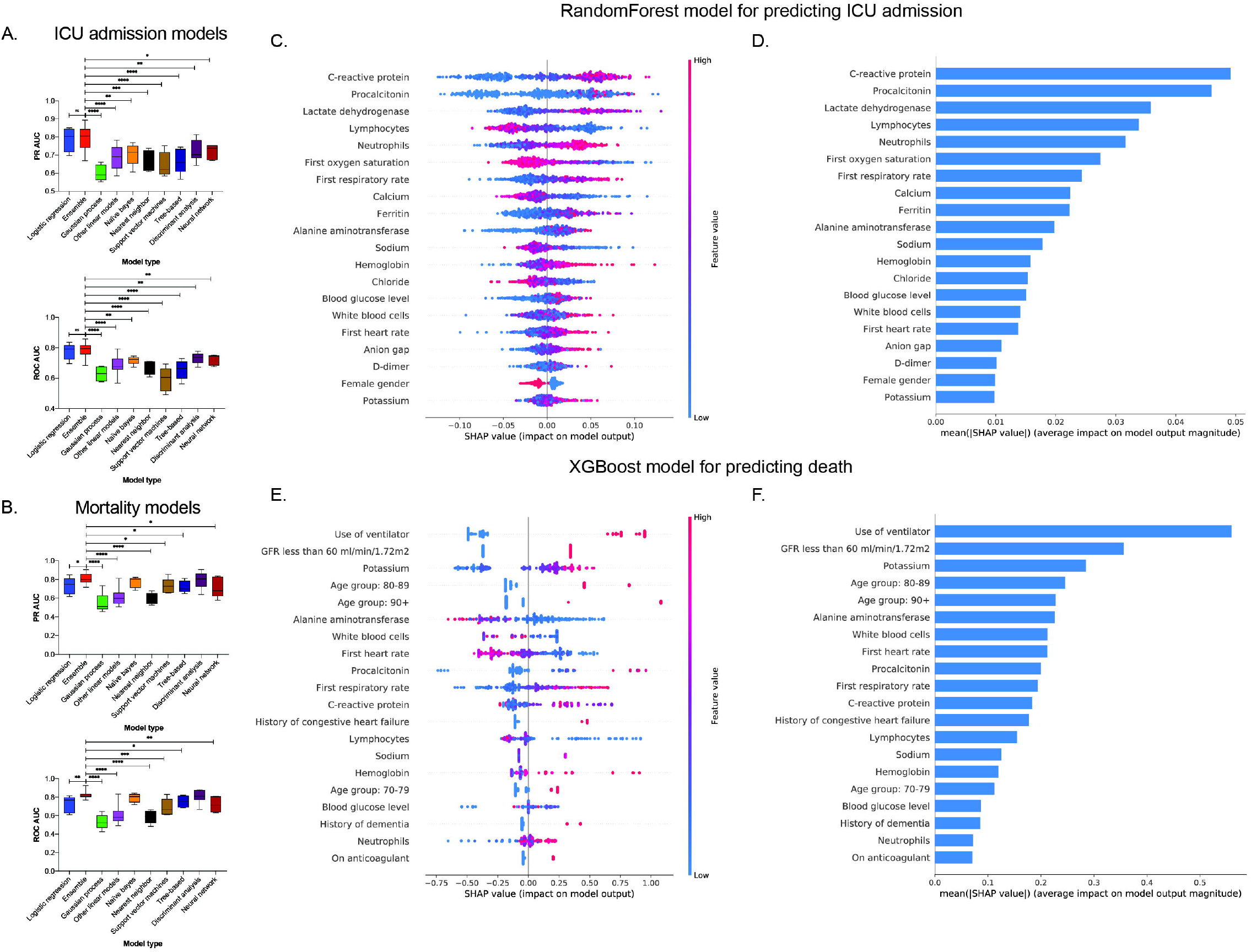
**(A-B). Boxplots** representing the precision recall area under the curve (PR AUC) and receiver operating characteristic area under the curve (ROC AUC) scores of ICU admission and mortality prediction models. Error bars indicate minimum and maximum values. Statistical analysis was performed using Fisher’s Least Significant Difference (LSD) *t*-test. *p-*value style is geometric progression - <0.03 (*), <0.002 (**), <0.0002 (***), <0.0001 (****). **Variables of importance for ICU admission and mortality prediction models**. (C) SHAP value summary dot plot and (D) variable of importance of *RandomForest* algorithm-based ICU admission model. (E) SHAP value summary dot plot and (F) variable of importance of *XGBClassifier* algorithm-based mortality model. The calculation of SHAP values is done by comparing the prediction of the model with and without the feature in every possible way of adding the feature to the model. The bar plot depicts the mean SHAP values whereas the summary dot plot shows the impact on the model. The color of the dot represents the value of the feature and the X-axis depicts the direction and magnitude of the impact. Red colored dots represent high value of the feature and the blue represents lower value. A positive SHAP value means the feature value increases likelihood of ICU admission/mortality. For features with positive SHAP value for red dots, suggests directly proportional variable to outcome of interest and those with positive SHAP value for blue dots, suggest inverse correlation.

On comparing the mortality prediction models using cross validation, all ensemble-based models had mean PR AUC scores higher than 0.8 (Table 3; Fig. S2C-D). The PR AUC score for *AdaBoostClassifier* was 0.81 (95% CI, 0.76 – 0.86), for *BaggingClassifier* was 0.81 (95% CI, – 0.88), for *GradientBoostingClassifier* was 0.81 (95% CI, 0.73 – 0.89), for *RandomForestClassifier* was 0.8 (95% CI, 0.75 – 0.85), for *XGBClassifier* was 0.82 (95% CI, – 0.89), and ExtraTreesClassifier [0.82 (95% CI, 0.74 – 0.90)]. In addition, *LinearDiscriminantAnalysis* [0.85 (95% CI, 0.79 – 0.91)] also had a high PR AUC score. However, for mortality prediction, *LogisticRegression* [0.73 (95% CI, 0.62 – 0.84)] had low PR AUC score when compared to ensemble methods. The lowest PR AUC scores were for *GaussianProcessClassifier* [0.55 (95% CI, 0.42 – 0.68)], *SGDClassifier* [0.54 (95% CI, 0.49 – 0.59)], Perceptron [0.6 (95% CI, 0.53 – 0.67)], and *KNeighborsClassifier* [0.6 (95% CI, 0.52 – 0.68)]. Upon performing multiple comparison analysis between all models (based on PR AUC and ROC AUC scores), the ensemble-based models and *LinearDiscriminantAnalysis* models had similar patterns of performance (Fig. S1C-D). When we grouped the models based on their broad categories and compared their PR AUC and ROC AUC scores, we found that ensemble-based models perform better than all other model types except Naïve bayes and discriminant analysis based methods (based on Fisher’s Least Significant Difference (LSD) t-test; Fig. 2B; details of statistical analysis in Table S7).

**Table 3.**
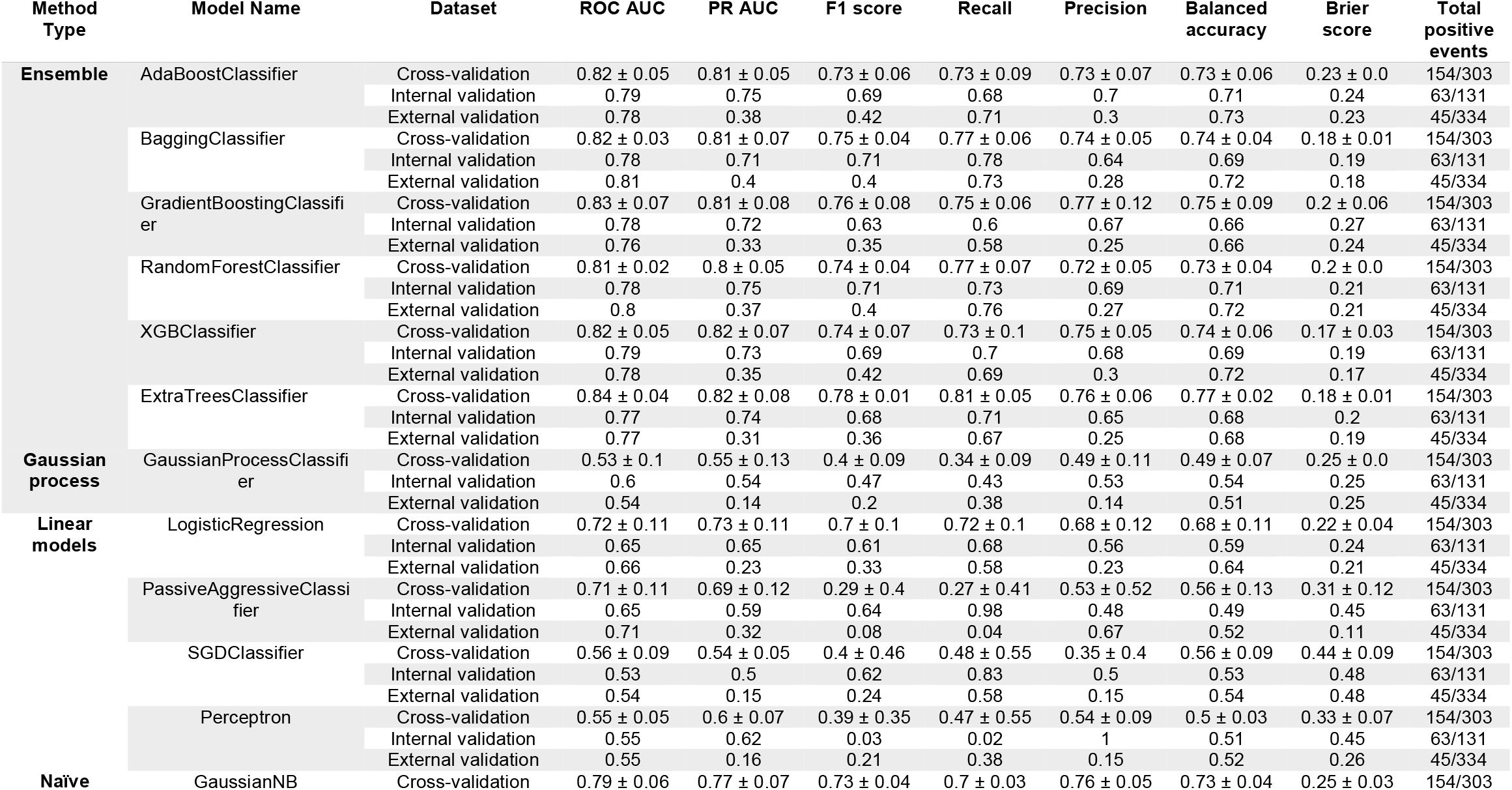
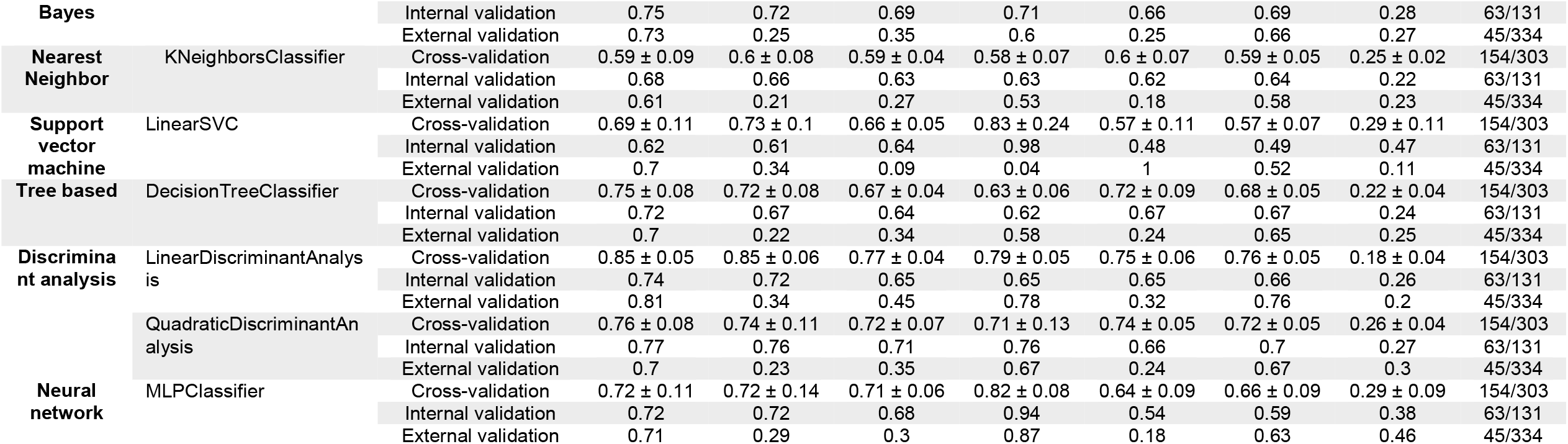
Performance of machine learning models to predict mortality within 28 days in COVID-19 patients. Cross-validation scores are expressed as mean ± 95% confidence interval.

### Comparing performance of prediction models – internal and external validation

We then tested the internal validation dataset on ICU admission models and found that ensemble methods (PR AUC ≥ 0.8) and *LogisticRegression* (PR AUC = 0.83) had the best scores (Table 2). However, for the external validation dataset, *BaggingClassifier, RandomForestClassifier* and *XGBClassifier* had better PR AUC scores (≥ 0.6) than other ensemble models. *LogisticRegression* also performed comparably (PR AUC = 0.62) to well-performing ensemble methods with the external validation dataset.

On evaluating the performance of mortality models using internal validation dataset, ensemble methods, naïve bayes, and discriminant analysis-based models outperformed other models (PR AUC ≥ 0.7) (Table 3). In the external validation dataset, although the PR AUC scores were lower, *AdaBoostClassifier, BaggingClassifier*, and *RandomForestClassifier* had better PR AUC scores (≥ 0.37) than other models. Unlike ICU admission prediction, *LogisticRegression* had a low score with internal and external validation datasets (PR AUC = 0.65 and 0.23, respectively).

Overall, we found that ensemble models performed well in predicting both ICU admission and mortality for COVID-19 patients.

### Critical variables for predicting ICU admission and mortality

To investigate how individual variables in the machine learning models impact outcome prediction, we performed SHAP analysis for the best models – namely random forest for the ICU admission model and XGB classifier for the mortality prediction model. For the ICU admission prediction models, C-reactive protein, procalcitonin, lactate dehydrogenase, and first respiratory rate were directly proportional to risk of ICU admission (Fig. 2C-D), while lower values of the first oxygen saturation reading and lymphocytes were associated with increased probability of ICU admission. For mortality prediction models, use of ventilator, estimated glomerular filtration rate less than 60 ml/min/1.72 m^2^, age greater than 80 years, hyperkalemia and high procalcitonin were associated with higher mortality while lower lymphocyte counts were associated with increased probability of death (Fig. 2E-F).

## Discussion

In this study, we evaluated the ability of various machine learning algorithms to predict clinical outcomes such as ICU admission or mortality using data available from initial ER encounter of COVID-19 patients. Based on our analysis of 18 algorithms, we found that ensemble-based methods have moderately better performance than other machine learning algorithms. Optimizing the hyperparameters (Tables S4 and S5) enabled us to achieve the best-performing ensemble models. We also identified variables that had the largest impact on the performance of the models. We demonstrated that for predicting ICU admission, C-reactive protein, LDH, procalcitonin, lymphocytes, neutrophils, oxygen saturation and respiratory rate are among the top predictors, but for mortality prediction, eGFR < 60 ml/min/1.73m^2^, serum potassium levels, use of ventilator, age, ALT and white blood cells are the leading predictors.

Our model detected that CRP, LDH, procalcitonin, eGFR< 60 ml/min/m2, serum potassium levels, advanced age and ventilator use are indicative of a worse outcome, which aligns with previous studies of ICU admission and mortality (Table S2). Multiple retrospective studies showed that increased procalcitonin values were associated with high risk for severe COVID-19 infection^12^. The explanation behind this association is not clear. Increased procalcitonin level in COVID -19 patients can suggest bacterial coinfection, a marker of severity of ARDS and immune dysregulation^13-15^ but may also be a marker of the hyperinflammation associated with COVID-19 severity. We also found reduced kidney function as the major risk factor for ICU mortality. This result has been revealed by two previous studies in the literature, indicating that patients on dialysis and with chronic kidney disease have a high risk of mortality from COVID-19^16,17^. Our study also highlighted serum potassium level as an important predictor for mortality. This finding has not been reported in the literature to our knowledge, although one study has reported the high prevalence of hypokalemia among patients with COVID-19^18^. Potassium derangement is independently associated with increased mortality in ICU patients^19^. Deviations in serum potassium levels in COVID-19 patients may suggest dysregulation of the renin-angiotensin system^20^ which has been suggested to also play a role in SARS-CoV-2 pathogenesis. This finding shows that the model aligns with previously reported clinically relevant markers and also predicts new markers that emerged from our patient population.

Our study utilizes a multi-hospital cohort that has been developed and validated in temporarily distinct subsets of the cohort. Multiple studies in the past using machine learning methodology to study COVID-19 outcomes used only a few machine learning algorithms^8-10,21,22^. However, these studies were oriented toward identifying clinical features rather than determining the best machine learning algorithm at predicting clinical outcomes, so only limited number of models were tested. To our knowledge, this is the first study to quantitatively and systematically compare 18 machine learning models through robust methodology encompassing all categories of algorithms. We showed that ensemble-methods perform better than other methods in predicting ICU admission and mortality from COVID-19. Ensemble methods are meta-algorithms that combine several different machine learning techniques into one unified predictive model (Table S3)^23^, which could explain this improvement in performance. We have also done exhaustive hyperparameter tuning to determine the best values. By performing SHAP analysis, we showed how variables impact outcomes in black-box machine learning models. Thus, our study is consistent with previous clinical study results, revealing similar clinical predictors for ICU admission and mortality, utilizing higher-performing machine learning models.

There are a number of limitations in our study. The lack of complete laboratory values for all patients necessitated exclusion of a large number of patients and removal of some variables in development of the models. As suggested by Jakobsen et al^24^, imputation is not an advisable method to handle missingness, when the percentage of missing data exceeds 40%. The majority of individuals (>98%) included in our analysis were those patients who visited to ED and subsequently became in-patients. In the patients excluded due to missingness, only ∼40% of the patients needed in-patient care. This discrepancy in severity might be the reason for lack of laboratory values in excluded patients.

Another limitation is that, as some of the laboratory values may take hours to be reported, the data may not be available until after the patient has transitioned out of the ER, decreasing the utility of using these predictors in triaging patient disposition. Similarly, as the mortality model uses ventilator use as a predictor, it requires ICU admission to be utilized and would not be valid in an earlier phase of care.

We also observed that the predicting capability on the external cohort (imbalanced dataset) was higher for ICU admission models in comparison to mortality models. This could be due to the changes instated in the ICU during the later period of pandemic. The changes in the treatment regimens might be affecting the mortality and thereby affecting the predictive power of our models. Our cohort is based on the population from Southern New England region of United States and includes two hospitals that are world-class academic centers, which could also limit the versatility of the models. More elaborate studies based on this framework in other cohorts would help validate our findings.

Our model development process and findings could be used by clinicians in gauging the clinical course, particularly ICU admission, of an individual with COVID-19 during an ED encounter. We would recommend using ensemble-based methods for developing clinical prediction models. Our ensemble methods identified key features in patients, such as kidney function, potassium, procalcitonin, CRP and LDH, that allowed us to predict clinical outcomes. Deploying such models would augment the clinical decision-making process by allowing physicians to identify potentially high-risk individuals and adjust their treatment accordingly.

## Methods

### Study population

Patients from the Mass General Brigham (MGB) healthcare system that were positive for COVID-19 between March and August of 2020 and had an ED encounter were included. Patients either had COVID-19 prior to the index ED visit or were diagnosed during that encounter. MGB is an integrated health care system which encompasses 14 hospitals across the New England area in the United States. COVID-19 positive patients were defined by the COVID-19 infection status, a discretely recorded field in the Epic EHR (Epic, Inc., Verona, WI). The COVID-19 infection status was added automatically if a SARS-CoV-2 PCR test was positive, or by Infection Control personnel if the patient has a confirmed positive test from an outside facility. This study was approved by the MGB Institutional Review Board.

### Data collection and covariate selection

We queried the data warehouse of our EHR for patient-level data including demographics, comorbidities, home medications, most recent outpatient recorded blood pressure, and death date. For each hospital encounter we extracted vital signs, laboratory values, admitting service, hospital length of stay, date of first ICU admission, amongst others. The patient’s problem list was extracted and transformed into a comorbidity matrix by using the “comorbidity” R package ^25^.

### Outcome definition

The two primary outcomes used for developing the models were ICU admission within 5 days of ED encounter and mortality within 28 days of ED encounter. The beginning of the prediction window began upon arrival to the ED.

### Model development

As described in Table S1, we selected a reduced set of potential predictor variables from previously published literature (Table S2). We used the same covariates in developing the ICU admission and mortality models except for ventilator use which was added to mortality models but excluded from ICU admission models. Age (10 year intervals), race (African American or other), BMI, modified Charlson Comorbidity Index ^26^, angiotensin converting enzyme inhibitor/angiotensin receptor blocker (ACEi/ARB) use, hypertension (>140/90 mmHg), and eGFR <60 ml/min were treated as categorical values. Patients with missing values for the independent variables or obviously incorrect entries (e.g., one patient was listed with respiratory rate of 75 breaths per minute) were excluded. Imputation was not advisable due to a high percentage of missingness^24^. Models were developed using the patients admitted during the period of March and April 2020. For external validation, we used a temporally distinct cohort consisting of patients admitted from May through August 2020. The data set was imbalanced with significantly fewer patients who were admitted to the ICU or died due to COVID-19 compared with those who did not. For the purpose of developing and internally validating the machine learning models, we randomly selected surviving patients who were not admitted to the ICU and matched the number of patients who were admitted to the ICU or died (n = 684 for ICU models and n = 434 for mortality models). From this group of patients, 70% (n = 478 for ICU models and n = 303 for mortality models) were used for developing machine learning models and the remaining 30% were used for internal validation.

A total of eighteen machine learning algorithms were tested, the descriptions of which are available in Table S3. For every machine learning model, we used a three-step approach. First, we made models using various combinations of tunable hyperparameters which are used to control the learning process of algorithms. The hyperparameters that were adjusted depended on the algorithm (outlined in Table S4). After developing these models for each combination of hyperparameter, we tested the performance of each of these combinations using a cross validation technique (number of folds = 5) during which a precision-recall area under curve (PR AUC) score was considered to select the best hyperparameter (Table S5). PR AUC score compares the positive predictive value (precision) and true positive rate (sensitivity or recall) of a model. For grading the performance of models, we used PR AUC scores as this is more applicable for datasets that are imbalanced. In our case, the external validation dataset remained an imbalanced dataset.

### Evaluation of model performance

Model performance evaluation was done in three parts. A *StratifiedKFold* technique of cross validation was first used during model development. In this method, 20% of the patients were excluded while training the model and the excluded patients were then used to test the model. This was done in an iterative process. Each model was evaluated by calculating the Receiver Operating Characteristic Area Under the Curve (ROC AUC), PR AUC, F1, recall, precision, balanced accuracy, and Brier scores. To calculate the 95% confidence interval, we used t_0.975, df = 4_ *=* 2.776 based on *t*-distribution for n = 5.

For the second level of validation, the model performance was evaluated on the 30% of patients who were not used during development of the models. This cohort worked as an internal validation dataset for these models. Finally, for the external validation, the cohort of patients who presented to the ED between May and August 2020 was used (Table S6).

### Model interpretation using Shapley values

For explaining the models, SHAP feature importance was reported based on Shapley values ^27^, details of which are outlined in the Supplementary Methods. SHAP values are useful to explain “black-box” machine learning models which are otherwise difficult to interpret. SHAP values for each patient feature explain the intensity and direction of impact on predicting the outcome.

#### Software

Data cleaning and processing were performed with R (R Core Team, version 3.6.3) using the tidyverse and comorbidity packages ^25,28,29^. Machine learning model development was done using Python (details in Supplementary Methods) ^30-33^. The programming code for R and Python are available upon request addressed to the corresponding author.

## Data Availability

The programming code for R and Python are available upon request addressed to the corresponding author.

## Supplementary Materials

### Methods

Fig. S1. Matrix plots showing differential model performance

Fig. S2. ROC AUC and PR AUC plots

Table S1. Selection of patients and variable details used for developing and testing the models

Table S2. Risk factors identified for mortality and ICU admission in COVID-19 studies

Table S3. Description of machine learning algorithms

Table S4. Hyperparameters which were optimized for machine learning algorithms

Table S5. Best hyperparameter values for machine learning algorithms that were chosen after tuning hyperparameters using *GridSearchCV* and *cross validation* technique.

Table S6. Characteristics of patients who visited the emergency room between May and August 2020 for COVID-19, that were used to evaluate the machine learning models as an external dataset. Variables stratified based on ICU admission and death of patients.

Table S7. Multiple comparison between ensemble methods and other types of machine learning algorithms using Fischer Least Significant Difference (LSD) t-test.

## Acknowledgments

Rakesh Jain’s research is supported by R01-CA208205, and U01-CA 224348, Outstanding Investigator Award R35-CA197743 and grants from the National Foundation for Cancer Research, Jane’s Trust Foundation, American Medical Research Foundation and Harvard Ludwig Cancer Center. We would like to thank Ashwin Srinivasan Kumar, Avanish Ranjan, Tariq Anwar and Mushtaq Rizvi for advice on machine learning algorithms and Python coding.

## Author contributions

S.S. performed, designed and built machine learning models. S.D. extracted data from the MGB database. A.V., A.B.P., C.C.H., M.J.K., S.D., H.L., T.S., L.L.M., and R.K.J supervised model development. All authors were involved in writing the article.

