## Supplementary Materials for "Comparing Machine Learning Algorithms for Predicting ICU Admission and Mortality in COVID-19"

**Methods**

***Model interpretation using Shapley values***

For explaining the models, SHAP feature importance was reported based on Shapley values. *TreeExplainer* based SHAP values were used to demonstrate the impact of covariates on model prediction. SHAP analysis was performed for best performing model for ICU admission and mortality prediction.

***Softwares and packages***

For processing the clinical data, dplyr and tableone packages in R were used. After processing, model development was performed in Python (version 3.8; Python Software Foundation) using open-source packages including *sklearn*, *xgboost*, *imblearn*, *matplotlib* and *shap* packages. *GridsearchCV* and *cross_val_score* functions were used to identify the best hyperparameters. For comparing model types, GraphPad prism software (version 8.4.3) was used ^30-33^.

Fig. S1. Matrix plots showing differential model performance

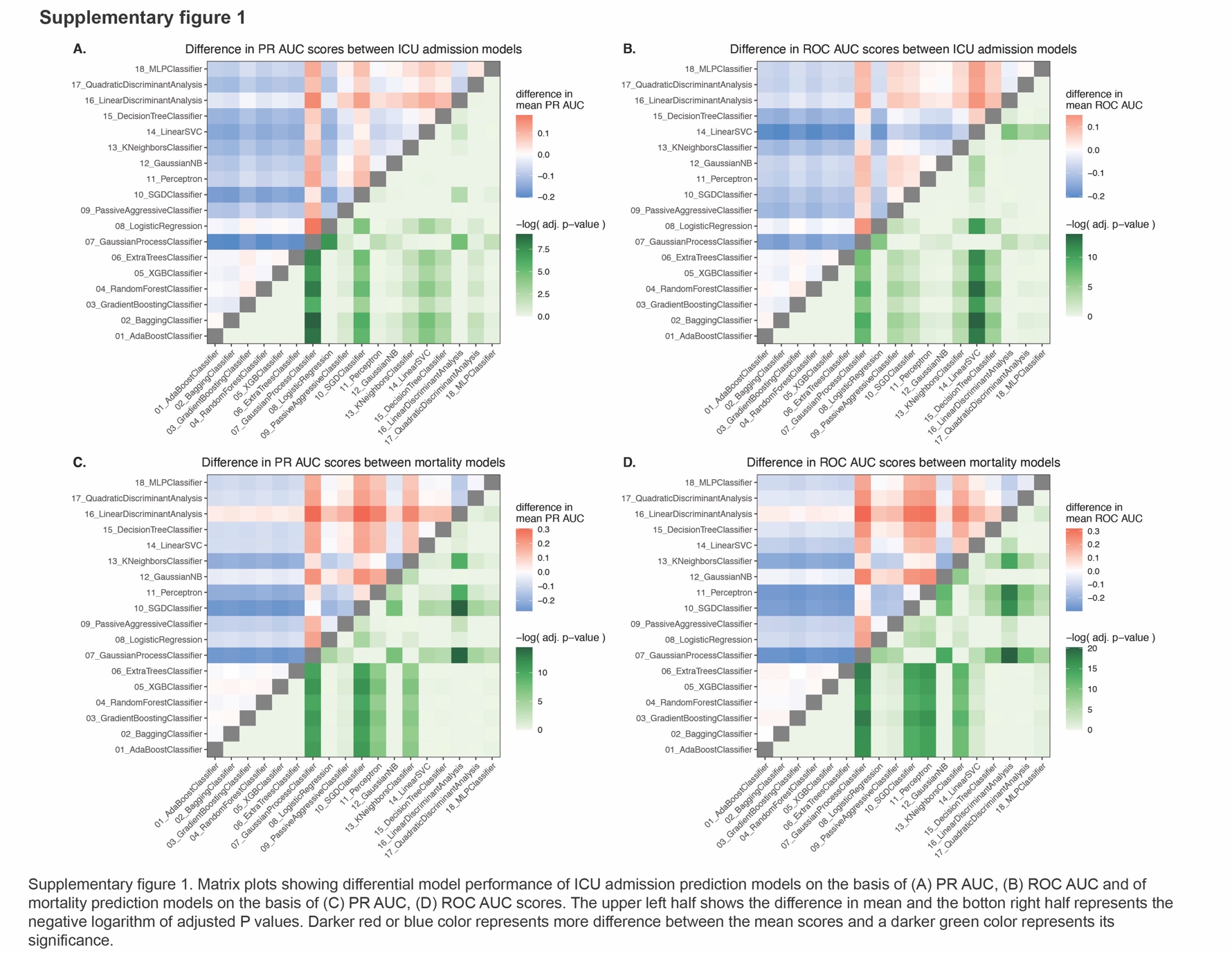

**Fig. S2.** ROC AUC and PR AUC plots

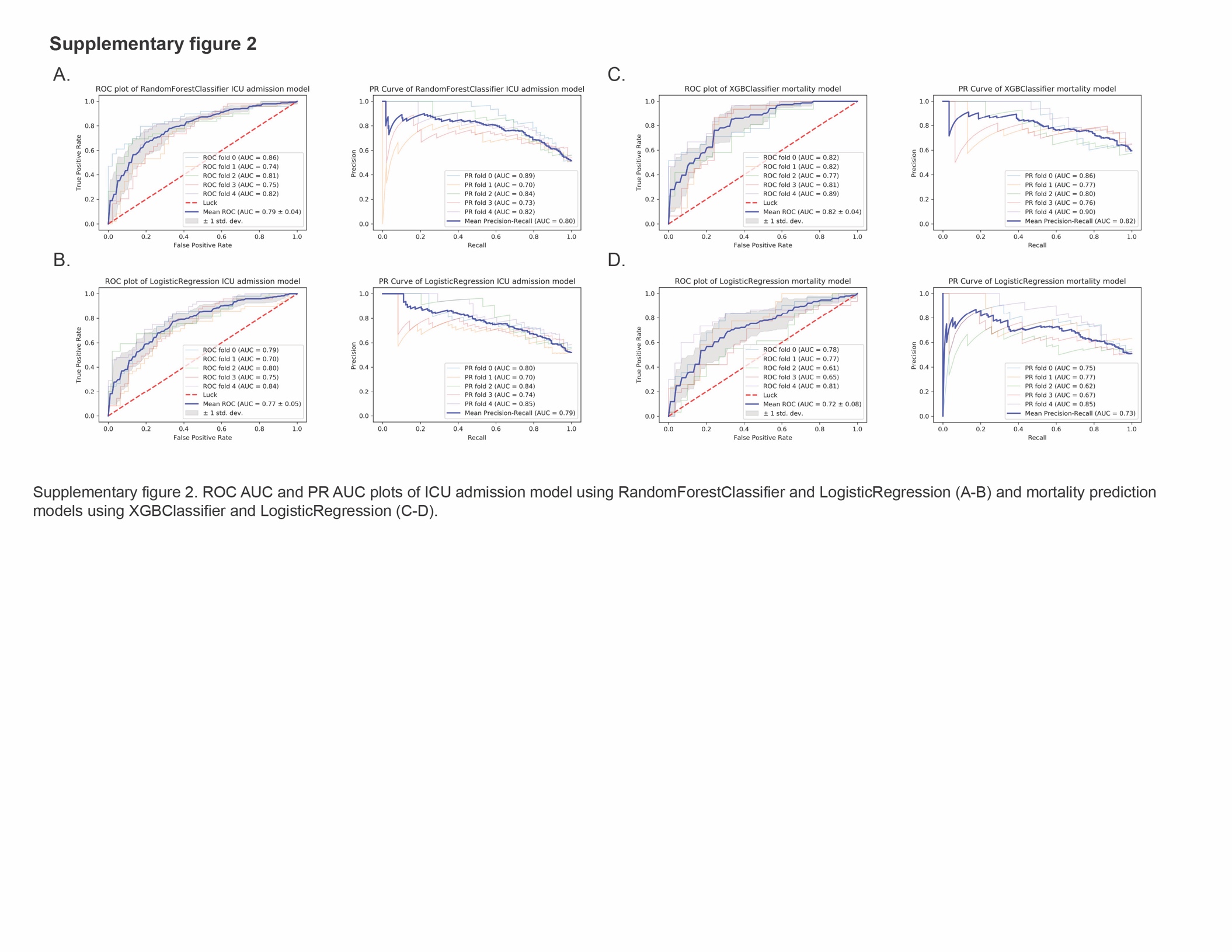

**Table S1.** Selection of patients and variable details used for developing and testing the models

|  | Dataset used for model development | Dataset used for external validation |
| --- | --- | --- |
|  | **March and April 2020** | **May to August 2020** |
| Total number of patients with COVID-19 in multi-hospital Mass General Brigham database | 10826 | 8013 |
| Number of patients which visited emergency department | 3713 | 1754 |
| Number of patients remaining after removing missingness in variables considered for model | 1144 | 334 |
| Number of patients included in the balanced dataset which was used for model development | 684 for ICU admission model and 434 for death prediction model | All 334 patients were tested - unbalanced dataset |
| Variable category | **Variable used for model development and testing** | |
| Demographic | age, sex, race, BMI | |
| Medication use | ACEi/ARB, ARA, Calcium channel blocker, Betablocker, vasodilator, alphablocker, diuretic, antiplatelet, NSAID, Proton pump inhibitor, statin, anticoagulant | |
| History of past illness | acute myocardial infarction, congestive heart failure, peripheral vascular disease, cerebrovascular disease, dementia, chronic obstructive pulmonary disease, rheumatic disease, peptic ulcer disease, mild liver disease, diabetes with or without complications, hemiplegia, renal disease, cancer, moderate/severe liver disease, metastatic cancer, AIDS, hypertension | |
| Clinical examination | First respiratory rate, first heart rate, first oxygen saturation | |
| Laboratory values | C-reactive protein, lymphocyte count, WBC count, neutrophil count, D-dimer, total bilirubin, lactate dehydrogenase, GFR (chronic kidney disease), anion gap, hemoglobin, procalcitonin, glucose, alanine aminotransferase, serum sodium, calcium, magnesium, potassium, chloride and ferritin | |

**Table S2.** Risk factors identified for mortality and ICU admission in COVID-19 studies

| Study authors | Study design | Period | Sample size | Risk factors for Mortality | Risk factor for ICU admission |
| --- | --- | --- | --- | --- | --- |
| Elizabeth J Williamson et al.(*16*) | Prospective cohort study | February -May 2020 | 17,278,392 | Male - (HR) 1.59 (1.53-1.65),  Black - (HR) 1.48 (1.29-1.69),  BMI > 40- (HR) 1.92(1.72-2.13),  eGFR< 60- (HR) 1.33(1.28-1.40),  Diabetes- (HR) 1.90(1.72-1.96),  Chronic heart disease- (HR) 1.17(1.12-1.22),  Cancer < 1 year ago- (HR) 1.72 (1.50-1.96) |  |
| Christopher M Petrilli et al.(*34*) | Prospective cohort study | March- April 2020 | 5279 | NA | Heart failure- (OR) 1.9 (1.4-2.5),  BMI> 40- (OR) 1.5 (1.0-2.2),  Male Sex- (OR) 1.5 (1.3-1.8),  Oxygen saturation < 88% -(OR) 3.7 (2.8-4.8),  Troponin level > 1 (OR) 4.8(2.1-10.9),  CRP > 200- (OR) 5.1(2.8-9.2),  D-dimer > 2500-  (OR) 3.9 (2.6-6.0) |
| Michaela R Anderson et al.(*35*) | Retrospective study | March – April  2020 | 2466 | BMI > 40 -(HR) 1.6 (1.1-2.1) | NA |
| Sara Y Tartof et al.(*36*) | Retrospective  study | February- May  2020 | 6916 | BMI 40-45 -(HR) 2.68 (1.43- 5.04),  BMI > 45- (HR) 4.18( 2.12-8.26) | NA |
| Matthew J Cummings et al.(*37*) | Prospective cohort study | March- April 2020 | 1150 | Older age- (HR) 1.31 (1.09-1.57) per 10 years increase,  Chronic cardiac disease – (HR) 1.76 (1.08-2.86),  Chronic pulmonary disease – (HR) 2.94 (1.48-5.84),  Higher concentrations of interleukin-6- (HR) 1.11 ( 1.02-1.20) per decile increase,  Higher concentrations of D-dimer- (HR) 1.11 (1.01-1.19) per decile increase |  |
| Shruti Gupta et al.(*38*) | Retrospective cohort study | March -April 2020 | 2215 | Older age (≥ 80 vs < 40 years- (OR) 11.15 (6.19-20.06),  Coronary artery disease- (OR) 1.47 (1.07-2.02),  Male- (OR) 1.50 (1.19-1.90),  BMI(≥ 40 vs < 25)- (OR) 1.51 (1.01-2.25),  Active cancer -(OR) 2.15 (1.35-3.43),  Pao2:Fio2 < vs ≥ 300 mm of Hg- (OR) 2.94 (2.11-4.08) | NA |
| Giacomo Grasselli et al.(*39*) | Retrospective cohort study | February- April 2020 | 3988 | Older age -(HR) 1.75 (1.60-1.92),  Male sex- (HR) 1.57 (1.31-1.88),  High Fio2- (HR) 1.14 (1.10-1.19),  High PEEP (HR) 1.04 (1.01-1.06)  Type 2 DM-(HR) 1.18 (1.01-1.39) | NA |
| Jonathan W.Cunningham et al.(*40*) | Retrospective study | April -June 2020 | 63103 | Morbid obesity- (OR) 2.30 (1.77-2.98),  Hypertension- (OR) 2.36 (1.79-3.12),  Male sex- (OR) 1.53 (1.20-1.95) | NA |

**Table S3.** Description of machine learning algorithms

| Model Type | Model name | Description |
| --- | --- | --- |
| Ensemble | AdaBoostClassifier | It is a decision tree-based classifier where instead of a full tree, each tree only has one split, like a stump. Sequential ensembling is performed by boosting technique where an error that the first stump makes does impact the second stump. By fitting consecutive trees at every step, the accuracy increases than the previous tree. The drawback, however, is the chance of overfitting. |
|  | BaggingClassifier | The bagging classifier fits base classifiers (default is decision tree) on random subsets of the training dataset and then average the individual predictions based on voting to form a final prediction. Here all variables are considered for splitting a node. |
|  | GradientBoostingClassifier | In this machine learning technique, multiple weak prediction models are combined to form more robust prediction model. The negative gradient of the loss function (linked with the whole ensemble) is maximally correlated with weak learners. |
|  | RandomForestClassifier | It is a decision tree-based classifier, which runs multiple trees in parallel. It uses a bagging technique to reduce variance, in which numerous subsets of data from the training sample are chosen randomly with replacement to train decision trees. The average of all predictions from decision trees is considered, which also reduces over-fitting. Unlike bagging classifier, here only a subset of features is considered at random and the best split is used. |
|  | XGBClassifier | Extreme gradient boosting (XGB) classifier is similar to gradient boosting classifier, except that it uses second-order gradients of the loss function and uses advanced regularization, which improved model generalization. It is computed in parallel and also trains fast. |
|  | ExtraTreesClassifier | It is an algorithm similar to Random forest classifier, where a subset of features is used but instead of considering the most discriminative threshold, randomly drawn thresholds are used. The best randomly generated threshold is then picked as the splitting rule. |
| Gaussian process | GaussianProcessClassifier | It implements Gaussian processes for probabilistic classification. It uses a latent function which allows a convenient formulation of model which is then removed during prediction. |
| Linear models | LogisticRegression | It is a machine learning algorithm in which the dependent categorical variable is predicted as a function of independent variables. Here regularization technique was used to prevent over-fitting. |
|  | PassiveAggressiveClassifier | It is a linear model in which if prediction is correct, the model estimates are kept same but if prediction is incorrect, alterations are made in the model. They do not have a learning rate but they do have a regularization hyperparameter. |
|  | SGDClassifier | It is a linear classifier that uses stochastic gradient descent as a solver and allows the usage a numerous loss function. |
|  | Perceptron | It is also a linear model with stochastic gradient descent but is uses a set of functions to learn from inputs to ascertain if they belong to one class or the other. |
| Naïve Bayes | GaussianNB | It is a supervised machine learning algorithm which is based upon bayes theorem and it assumes that every feature is independent from values of other features for a given class. |
| Nearest Neighbor | KNeighborsClassifier | It is a machine learning algorithm where a user defined constant is considered for calculating the number of neighbors that can be grouped into one class. Here the distance between samples are generally calculated in terms of standard Euclidean distance. |
| Support vector machine | LinearSVC | It is a supervised machine learning algorithm where training data is fit to a model in which best fit hyperplanes divide the data into outcome categories. |
| Tree based | DecisionTreeClassifier | It is a non-parametric supervised machine learning method where a model is made to predict target variable by learning simple decision rules from the features provided in the training data. |
| Discriminant analysis | LinearDiscriminantAnalysis | It is a classifier algorithm with a linear decision boundary. The model is made by fitting class conditional densities to the training data. Gaussian distribution and similar covariance are the assumptions used by this algorithm. |
|  | QuadraticDiscriminantAnalysis | It is a machine learning algorithm similar to Linear Discriminant analysis but with a quadratic decision boundary. |
| Neural network | MLPClassifier | MLPClassifier (multilayer perceptron classifier) is a type of neural network-based machine learning algorithm where the layers are formed using multiple layers (atleast three layers) of perceptron. |

**Table S4.** Hyperparameters which were optimized for machine learning algorithms

| Model Name | Hyperparameters optimized |
| --- | --- |
| AdaBoostClassifier' | 'n_estimators': [10, 100, 1000, 10000],  'learning_rate': [0.001, 0.01, 0.1, 1] |
| BaggingClassifier' | 'n_estimators': [10, 100, 1000, 10000],  'max_samples': [1, 10, 100, 1000] |
| GradientBoostingClassifier' | 'n_estimators': [10, 100, 1000, 10000],  'learning_rate': [0.001, 0.01, 0.1, 1] |
| RandomForestClassifier' | 'n_estimators': [10, 100, 1000, 10000],  'max_features': ['auto', 'log2'],  'max_depth': [2, 5, 10, 20, 50, 100],  'criterion': ['gini', 'entropy'] |
| XGBClassifier' | 'max_depth': [2, 5, 10, 20, 50, 100],  'n_estimators': [10, 100, 1000, 10000],  'learning_rate': [0.001, 0.01, 0.1, 1] |
| ExtraTreesClassifier' | 'n_estimators': [100, 1000, 10000],  'criterion': ['gini', 'entropy'],  'max_features': ['auto', 'log2'],  'max_depth': [2, 5, 10, 20, 50, 100] |
| GaussianProcessClassifier' | 'kernel': (RBF(length_scale=1), Matern(length_scale=1, nu=1.5), RationalQuadratic(alpha=1.5, length_scale=1)) |
| LogisticRegression' | 'penalty': ['l1', 'l2'],  'C': [0.1, 1, 10, 100, 200, 500, 1000] |
| PassiveAggressiveClassifier' | 'C': [0.0001, 0.0003, 0.001, 0.003, 0.01],  'loss': ['hinge', 'squared_hinge'],  'n_iter_no_change': [5, 10, 30, 100, 300] |
| SGDClassifier' | 'loss': ['modified_huber'],  'alpha': [0.01, 0.1, 0.5, 1],  'penalty': ['l2', 'l1', None] |
| Perceptron' | 'alpha': [0.0001, 0.001, 0.01],  'penalty': ['l2', 'l1', None] |
| GaussianNB' | NA |
| KNeighborsClassifier' | 'n_neighbors': [3, 5, 10, 20],  'leaf_size': [2, 5, 10, 20],  'p': [0.5, 1, 2, 5],  'weights': ['uniform', 'distance'],  'algorithm': ['auto', 'ball_tree', 'kd_tree', 'brute'] |
| LinearSVC' | 'penalty': ['l1', 'l2'],  'loss': ['hinge', 'squared_hinge'],  'C': [0.1, 0.5, 1, 5, 10] |
| DecisionTreeClassifier' | 'criterion': ['gini', 'entropy'],  'splitter': ['random', 'best'],  'max_depth': [1, 2, 10],  'min_samples_leaf': [1, 2, 10] |
| LinearDiscriminantAnalysis' | 'solver': ['svd', 'lsqr', 'eigen'],  'tol': [1e-05, 0.0001, 0.0003] |
| QuadraticDiscriminantAnalysis' | 'reg_param': [0.1, 0.5, 0.7, 0.9],  'tol': [1e-05, 0.0001, 0.0003] |
| MLPClassifier' | 'solver': ['lbfgs', 'adam'],  'learning_rate': ['constant', 'invscaling', 'adaptive'],  'hidden_layer_sizes': [(10, 7, 3), (30, 20, 12), (50, 35, 25), (70, 50, 35)],  'activation': ['identity', 'logistic', 'tanh', 'relu'] |

**Table S5.** Best hyperparameter values for machine learning algorithms that were chosen after tuning hyperparameters using *GridSearchCV* and *cross validation* technique.

| Method Type | Model name | Total fits tested | Best hyperparameter values for ICU admission prediction | Best hyperparameter values for death prediction |
| --- | --- | --- | --- | --- |
| Ensemble | AdaBoostClassifier | 80 | 'learning_rate': 0.01, 'n_estimators': 1000 | 'learning_rate': 0.001, 'n_estimators': 10000 |
|  | BaggingClassifier | 80 | 'max_samples': 10, 'n_estimators': 10000 | 'max_samples': 100, 'n_estimators': 10000 |
|  | GradientBoostingClassifier | 80 | 'learning_rate': 0.1, 'n_estimators': 100 | 'learning_rate': 0.1, 'n_estimators': 1000 |
|  | RandomForestClassifier | 480 | 'criterion': 'entropy', 'max_depth': 10, 'max_features': 'log2', 'n_estimators': 100 | 'criterion': 'gini', 'max_depth': 2, 'max_features': 'log2', 'n_estimators': 100 |
|  | XGBClassifier | 480 | 'learning_rate': 0.01, 'max_depth': 2, 'n_estimators': 1000 | 'learning_rate': 0.1, 'max_depth': 2, 'n_estimators': 100 |
|  | ExtraTreesClassifier | 360 | 'criterion': 'entropy', 'max_depth': 20, 'max_features': 'auto', 'n_estimators': 1000 | 'criterion': 'entropy', 'max_depth': 5, 'max_features': 'log2', 'n_estimators': 100 |
| Gaussian process | GaussianProcessClassifier | 15 | 'kernel': RationalQuadratic(alpha=1.5, length_scale=1) | 'kernel': RationalQuadratic(alpha=1.5, length_scale=1) |
| Linear models | LogisticRegression | 70 | 'C': 200, 'penalty': 'l2' | 'C': 100, 'penalty': 'l2' |
|  | PassiveAggressiveClassifier | 250 | 'C': 0.0001, 'loss': 'squared_hinge', 'n_iter_no_change': 10 | 'C': 0.0001, 'loss': 'hinge', 'n_iter_no_change': 100 |
|  | SGDClassifier | 60 | 'alpha': 0.1, 'loss': 'modified_huber', 'penalty': 'l1' | 'alpha': 0.01, 'loss': 'modified_huber', 'penalty': 'l1' |
|  | Perceptron | 45 | 'alpha': 0.01, 'penalty': 'l1' | 'alpha': 0.01, 'penalty': 'l1' |
| Naïve Bayes | GaussianNB | 5 |  |  |
| Nearest Neighbor | KNeighborsClassifier | 2560 | 'algorithm': 'auto', 'leaf_size': 2, 'n_neighbors': 20, 'p': 1, 'weights': 'distance' | 'algorithm': 'auto', 'leaf_size': 2, 'n_neighbors': 20, 'p': 1, 'weights': 'distance' |
| Support vector machine | LinearSVC | 100 | 'C': 1, 'loss': 'squared_hinge', 'penalty': 'l2' | 'C': 0.5, 'loss': 'squared_hinge', 'penalty': 'l2' |
| Tree based | DecisionTreeClassifier | 180 | 'criterion': 'entropy', 'max_depth': 10, 'min_samples_leaf': 10, 'splitter': 'random' | 'criterion': 'gini', 'max_depth': 10, 'min_samples_leaf': 10, 'splitter': 'random' |
| Discriminant analysis | LinearDiscriminantAnalysis | 45 | 'solver': 'svd', 'tol': 1e-05 | 'solver': 'svd', 'tol': 1e-05 |
|  | QuadraticDiscriminantAnalysis | 60 | 'reg_param': 0.9, 'tol': 1e-05 | 'reg_param': 0.1, 'tol': 1e-05 |
| Neural network | MLPClassifier | 480 | 'activation': 'logistic', 'hidden_layer_sizes': (30, 20, 12), 'learning_rate': 'constant', 'solver': 'adam' | 'activation': 'identity', 'hidden_layer_sizes': (10, 7, 3), 'learning_rate': 'constant', 'solver': 'adam' |

**Table S6.** Characteristics of patients who visited the emergency room between May and August 2020 for COVID-19, that were used to evaluate the machine learning models as an external dataset. Variables stratified based on ICU admission and death of patients.

|  |  | ICU admission | | | | | Death | | |
| --- | --- | --- | --- | --- | --- | --- | --- | --- | --- |
|  | Overall | | No | Yes | p | No | | Yes | p |
| n | 334 | | 260 | 74 | 0 | 289 | | 45 | 0 |
| Demographics |  | |  |  |  |  | |  |  |
| Age group (%) |  | |  |  | 0.33 |  | |  | <0.001 |
| 20-29 | 19 ( 5.7) | | 17 ( 6.5) | 2 ( 2.7) | 0 | 18 ( 6.2) | | 1 ( 2.2) | 0 |
| 30-39 | 26 ( 7.8) | | 23 ( 8.8) | 3 ( 4.1) | 0 | 26 ( 9.0) | | 0 ( 0.0) | 0 |
| 40-49 | 29 ( 8.7) | | 23 ( 8.8) | 6 ( 8.1) | 0 | 28 ( 9.7) | | 1 ( 2.2) | 0 |
| 50-59 | 58 (17.4) | | 45 (17.3) | 13 ( 17.6) | 0 | 55 (19.0) | | 3 ( 6.7) | 0 |
| 60-69 | 77 (23.1) | | 56 (21.5) | 21 ( 28.4) | 0 | 68 (23.5) | | 9 ( 20.0) | 0 |
| 70-79 | 43 (12.9) | | 29 (11.2) | 14 ( 18.9) | 0 | 36 (12.5) | | 7 ( 15.6) | 0 |
| 80-89 | 59 (17.7) | | 49 (18.8) | 10 ( 13.5) | 0 | 42 (14.5) | | 17 ( 37.8) | 0 |
| 90+ | 23 ( 6.9) | | 18 ( 6.9) | 5 ( 6.8) | 0 | 16 ( 5.5) | | 7 ( 15.6) | 0 |
| Sex = Male (%) | 185 (55.4) | | 146 (56.2) | 39 ( 52.7) | 0.693 | 159 (55.0) | | 26 ( 57.8) | 0.853 |
| Race = Other (%) | 275 (82.3) | | 218 (83.8) | 57 ( 77.0) | 0.236 | 236 (81.7) | | 39 ( 86.7) | 0.543 |
| BMI_categorical (%) |  | |  |  | 0.556 |  | |  | 0.033 |
| [0,25] | 98 (29.3) | | 80 (30.8) | 18 ( 24.3) | 0 | 80 (27.7) | | 18 ( 40.0) | 0 |
| (25,30] | 95 (28.4) | | 72 (27.7) | 23 ( 31.1) | 0 | 79 (27.3) | | 16 ( 35.6) | 0 |
| (30,75] | 141 (42.2) | | 108 (41.5) | 33 ( 44.6) | 0 | 130 (45.0) | | 11 ( 24.4) | 0 |
| Medication use |  | |  |  |  |  | |  |  |
| On ACEi/ARB = TRUE (%) | 78 (23.4) | | 62 (23.8) | 16 ( 21.6) | 0.808 | 67 (23.2) | | 11 ( 24.4) | 1 |
| On ARA = TRUE (%) | 8 ( 2.4) | | 8 ( 3.1) | 0 ( 0.0) | 0.273 | 6 ( 2.1) | | 2 ( 4.4) | 0.658 |
| On Calcium channel blocker = TRUE (%) | 53 (15.9) | | 35 (13.5) | 18 ( 24.3) | 0.038 | 44 (15.2) | | 9 ( 20.0) | 0.551 |
| On Betablocker = TRUE (%) | 83 (24.9) | | 64 (24.6) | 19 ( 25.7) | 0.973 | 67 (23.2) | | 16 ( 35.6) | 0.109 |
| On Vasodilator = TRUE (%) | 30 ( 9.0) | | 26 (10.0) | 4 ( 5.4) | 0.323 | 24 ( 8.3) | | 6 ( 13.3) | 0.414 |
| On Alphablocker = TRUE (%) | 9 ( 2.7) | | 8 ( 3.1) | 1 ( 1.4) | 0.688 | 8 ( 2.8) | | 1 ( 2.2) | 1 |
| On Diuretic = TRUE (%) | 76 (22.8) | | 66 (25.4) | 10 ( 13.5) | 0.046 | 64 (22.1) | | 12 ( 26.7) | 0.63 |
| On Antiplatelet = TRUE (%) | 12 ( 3.6) | | 12 ( 4.6) | 0 ( 0.0) | 0.126 | 10 ( 3.5) | | 2 ( 4.4) | 1 |
| On NSAIDs = TRUE (%) | 30 ( 9.0) | | 21 ( 8.1) | 9 ( 12.2) | 0.393 | 27 ( 9.3) | | 3 ( 6.7) | 0.761 |
| On Proton pump inhibitor = TRUE (%) | 86 (25.7) | | 70 (26.9) | 16 ( 21.6) | 0.442 | 70 (24.2) | | 16 ( 35.6) | 0.152 |
| On Statin = TRUE (%) | 127 (38.0) | | 100 (38.5) | 27 ( 36.5) | 0.863 | 104 (36.0) | | 23 ( 51.1) | 0.075 |
| On Anticoagulant = TRUE (%) | 42 (12.6) | | 37 (14.2) | 5 ( 6.8) | 0.13 | 32 (11.1) | | 10 ( 22.2) | 0.063 |
| History of past illness |  | |  |  |  |  | |  |  |
| Acute myocardial infarction = 1 (%) | 5 ( 1.5) | | 5 ( 1.9) | 0 ( 0.0) | 0.51 | 3 ( 1.0) | | 2 ( 4.4) | 0.275 |
| Congestive heart failure = 1 (%) | 42 (12.6) | | 34 (13.1) | 8 ( 10.8) | 0.749 | 31 (10.7) | | 11 ( 24.4) | 0.019 |
| Peripheral vascular disease = 1 (%) | 25 ( 7.5) | | 19 ( 7.3) | 6 ( 8.1) | 1 | 20 ( 6.9) | | 5 ( 11.1) | 0.491 |
| Cerebrovascular disease = 1 (%) | 40 (12.0) | | 33 (12.7) | 7 ( 9.5) | 0.58 | 30 (10.4) | | 10 ( 22.2) | 0.042 |
| Dementia = 1 (%) | 23 ( 6.9) | | 19 ( 7.3) | 4 ( 5.4) | 0.757 | 19 ( 6.6) | | 4 ( 8.9) | 0.8 |
| Chronic obstructive pulmonary disease = 1 (%) | 56 (16.8) | | 45 (17.3) | 11 ( 14.9) | 0.749 | 47 (16.3) | | 9 ( 20.0) | 0.682 |
| Rheumatic disease = 1 (%) | 2 ( 0.6) | | 1 ( 0.4) | 1 ( 1.4) | 0.923 | 2 ( 0.7) | | 0 ( 0.0) | 1 |
| Peptic ulcer disease = 1 (%) | 7 ( 2.1) | | 5 ( 1.9) | 2 ( 2.7) | 1 | 7 ( 2.4) | | 0 ( 0.0) | 0.62 |
| Mild liver disease = 1 (%) | 18 ( 5.4) | | 11 ( 4.2) | 7 ( 9.5) | 0.143 | 16 ( 5.5) | | 2 ( 4.4) | 1 |
| Diabetes = 1 (%) | 65 (19.5) | | 48 (18.5) | 17 ( 23.0) | 0.485 | 58 (20.1) | | 7 ( 15.6) | 0.611 |
| Diabetes with complications = 1 (%) | 18 ( 5.4) | | 13 ( 5.0) | 5 ( 6.8) | 0.765 | 14 ( 4.8) | | 4 ( 8.9) | 0.446 |
| Hemiplegia = 1 (%) | 5 ( 1.5) | | 4 ( 1.5) | 1 ( 1.4) | 1 | 4 ( 1.4) | | 1 ( 2.2) | 1 |
| Renal disease = 1 (%) | 49 (14.7) | | 37 (14.2) | 12 ( 16.2) | 0.811 | 37 (12.8) | | 12 ( 26.7) | 0.027 |
| Cancer = 1 (%) | 33 ( 9.9) | | 29 (11.2) | 4 ( 5.4) | 0.214 | 25 ( 8.7) | | 8 ( 17.8) | 0.101 |
| Moderate/severe liver disease = 1 (%) | 2 ( 0.6) | | 2 ( 0.8) | 0 ( 0.0) | 1 | 2 ( 0.7) | | 0 ( 0.0) | 1 |
| Metastatic cancer = 1 (%) | 1 ( 0.3) | | 0 ( 0.0) | 1 ( 1.4) | 0.502 | 0 ( 0.0) | | 1 ( 2.2) | 0.284 |
| AIDS = 1 (%) | 3 ( 0.9) | | 3 ( 1.2) | 0 ( 0.0) | 0.818 | 3 ( 1.0) | | 0 ( 0.0) | 1 |
| Hypertension = 1 (%) | 124 (37.1) | | 94 (36.2) | 30 ( 40.5) | 0.58 | 108 (37.4) | | 16 ( 35.6) | 0.945 |
| Laboratory values and clinical examination |  | |  |  |  |  | |  |  |
| CRP (mg/L) (mean (SD)) | 77.84 (80.07) | | 62.24 (64.07) | 132.65 (103.76) | <0.001 | 70.72 (76.35) | | 123.55 (88.85) | <0.001 |
| First respiratory rate (counts/min) (mean (SD)) | 22.46 (6.35) | | 21.33 (5.69) | 26.43 (6.94) | <0.001 | 22.08 (5.77) | | 24.87 (8.95) | 0.006 |
| First heart rate (beats/min) (mean (SD)) | 97.51 (21.27) | | 96.19 (20.33) | 102.16 (23.87) | 0.033 | 97.25 (21.62) | | 99.20 (18.97) | 0.569 |
| Sodium (mmol/L) (mean (SD)) | 137.31 (5.62) | | 137.55 (5.23) | 136.49 (6.78) | 0.153 | 137.09 (5.47) | | 138.76 (6.39) | 0.064 |
| Calcium (mg/dL) (mean (SD)) | 9.05 (0.62) | | 9.08 (0.60) | 8.96 (0.71) | 0.139 | 9.06 (0.62) | | 8.99 (0.63) | 0.467 |
| Magnesium (mg/dL) (mean (SD)) | 1.98 (0.31) | | 1.97 (0.28) | 2.01 (0.39) | 0.4 | 1.97 (0.29) | | 2.03 (0.41) | 0.215 |
| Potassium (mmol/L) (mean (SD)) | 4.16 (0.68) | | 4.12 (0.62) | 4.31 (0.85) | 0.031 | 4.12 (0.67) | | 4.37 (0.73) | 0.025 |
| Chloride (mmol/L) (mean (SD)) | 98.81 (5.54) | | 99.05 (5.35) | 97.97 (6.13) | 0.142 | 98.63 (5.24) | | 99.93 (7.16) | 0.144 |
| Lymphocytes (percentage; ref = 22-44%) (mean (SD)) | 18.34 (11.17) | | 19.44 (10.81) | 14.50 (11.62) | 0.001 | 19.00 (10.39) | | 14.09 (14.69) | 0.006 |
| Neutrophils (percentage; ref = 40-70%) (mean (SD)) | 70.83 (13.63) | | 69.34 (12.83) | 76.06 (15.11) | <0.001 | 70.04 (13.11) | | 75.92 (15.83) | 0.007 |
| WBC (x1000/µL) (mean (SD)) | 8.29 (5.52) | | 7.60 (4.34) | 10.75 (8.04) | <0.001 | 7.88 (4.40) | | 10.94 (9.80) | 0.001 |
| D-dimer (ng/mL) (mean (SD)) | 2635.76 (9620.95) | | 1864.46 (4388.44) | 5345.72 (18556.59) | 0.006 | 1818.14 (4181.74) | | 7886.67 (23526.20) | <0.001 |
| Total bilirubin (mg/dL) (mean (SD)) | 0.64 (1.27) | | 0.64 (1.38) | 0.63 (0.73) | 0.933 | 0.60 (0.90) | | 0.90 (2.61) | 0.133 |
| Ferritin (µg/L) (mean (SD)) | 698.76 (1012.15) | | 598.75 (796.45) | 1050.17 (1503.93) | 0.001 | 674.91 (974.35) | | 851.96 (1229.46) | 0.276 |
| LDH (Units) (mean (SD)) | 323.06 (171.78) | | 285.38 (115.15) | 455.45 (254.44) | <0.001 | 311.48 (159.87) | | 397.40 (222.45) | 0.002 |
| Low GFR (<60 ml/min/1.73m2) = TRUE (%) | 134 (40.1) | | 93 (35.8) | 41 ( 55.4) | 0.004 | 104 (36.0) | | 30 ( 66.7) | <0.001 |
| Anion gap (mmol/L) (mean (SD)) | 15.27 (3.84) | | 14.73 (2.96) | 17.18 (5.63) | <0.001 | 15.16 (3.85) | | 16.02 (3.79) | 0.16 |
| Hemoglobin (g/dL) (mean (SD)) | 12.68 (2.22) | | 12.71 (2.25) | 12.55 (2.12) | 0.582 | 12.77 (2.20) | | 12.09 (2.27) | 0.056 |
| First O2 saturation (%) (mean (SD)) | 94.66 (5.31) | | 95.68 (3.24) | 91.05 (8.64) | <0.001 | 95.07 (4.80) | | 92.00 (7.37) | <0.001 |
| ventilator_use = TRUE (%) | 50 (15.0) | | NA | NA | NA | 38 (13.1) | | 12 ( 26.7) | 0.032 |
| Procalcitonin (ng/ml) (mean (SD)) | 1.10 (6.33) | | 0.49 (1.85) | 3.28 (12.84) | 0.001 | 0.71 (2.85) | | 3.67 (15.58) | 0.003 |
| Glucose (mg/dL) (mean (SD)) | 159.48 (95.23) | | 147.64 (74.54) | 201.07 (139.30) | <0.001 | 157.12 (89.64) | | 174.60 (125.57) | 0.253 |
| ALT (IU/L) (mean (SD)) | 37.13 (38.79) | | 35.05 (36.79) | 44.42 (44.63) | 0.067 | 37.65 (39.73) | | 33.80 (32.28) | 0.537 |

**Table S7.** Multiple comparison between ensemble methods and other types of machine learning algorithms using Fischer Least Significant Difference (LSD) t-test.

| ICU admission models | | | | | |
| --- | --- | --- | --- | --- | --- |
| Multiple comparison of PR AUC scores | | | | | |
| Fisher's LSD | **Mean Diff.** | **95.00% CI of diff.** | **Significant?** | **Summary** | **Individual P Value** |
| Ensemble vs. Logistic regression | 0.005279 | -0.05256 to 0.06312 | No | ns | 0.8563 |
| Ensemble vs. Gaussian process | 0.1894 | 0.1315 to 0.2472 | Yes | **** | <0.0001 |
| Ensemble vs. Other linear models | 0.1056 | 0.06772 to 0.1435 | Yes | **** | <0.0001 |
| Ensemble vs. Naïve bayes | 0.08549 | 0.02764 to 0.1433 | Yes | ** | 0.0043 |
| Ensemble vs. Nearest neighbor | 0.1147 | 0.05685 to 0.1725 | Yes | *** | 0.0002 |
| Ensemble vs. Support vector machines | 0.1427 | 0.08482 to 0.2005 | Yes | **** | <0.0001 |
| Ensemble vs. Tree-based | 0.1223 | 0.06445 to 0.1801 | Yes | **** | <0.0001 |
| Ensemble vs. Discriminant analysis | 0.06747 | 0.02375 to 0.1112 | Yes | ** | 0.0029 |
| Ensemble vs. Neural network | 0.07261 | 0.01477 to 0.1304 | Yes | * | 0.0145 |
| Multiple comparison of ROC AUC scores | | | | | |
| Fisher's LSD | **Mean Diff.** | **95.00% CI of diff.** | **Significant?** | **Summary** | **Individual P Value** |
| Ensemble vs. Logistic regression | 0.01153 | -0.03482 to 0.05789 | No | ns | 0.6219 |
| Ensemble vs. Gaussian process | 0.1572 | 0.1109 to 0.2036 | Yes | **** | <0.0001 |
| Ensemble vs. Other linear models | 0.09872 | 0.06837 to 0.1291 | Yes | **** | <0.0001 |
| Ensemble vs. Naïve bayes | 0.06726 | 0.02090 to 0.1136 | Yes | ** | 0.005 |
| Ensemble vs. Nearest neighbor | 0.1133 | 0.06695 to 0.1597 | Yes | **** | <0.0001 |
| Ensemble vs. Support vector machines | 0.1933 | 0.1470 to 0.2397 | Yes | **** | <0.0001 |
| Ensemble vs. Tree-based | 0.1296 | 0.08329 to 0.1760 | Yes | **** | <0.0001 |
| Ensemble vs. Discriminant analysis | 0.05615 | 0.02110 to 0.09119 | Yes | ** | 0.002 |
| Ensemble vs. Neural network | 0.06348 | 0.01712 to 0.1098 | Yes | ** | 0.0079 |
| Mortality models | | | | | |
| Multiple comparison of PR AUC scores | | | | | |
| Fisher's LSD | **Mean Diff.** | **95.00% CI of diff.** | **Significant?** | **Summary** | **Individual P Value** |
| Ensemble vs. Logistic regression | 0.07725 | 0.004690 to 0.1498 | Yes | * | 0.0372 |
| Ensemble vs. Gaussian process | 0.262 | 0.1894 to 0.3345 | Yes | **** | <0.0001 |
| Ensemble vs. Other linear models | 0.1972 | 0.1497 to 0.2447 | Yes | **** | <0.0001 |
| Ensemble vs. Naïve bayes | 0.03971 | -0.03284 to 0.1123 | No | ns | 0.2793 |
| Ensemble vs. Nearest neighbor | 0.2072 | 0.1347 to 0.2798 | Yes | **** | <0.0001 |
| Ensemble vs. Support vector machines | 0.08152 | 0.008964 to 0.1541 | Yes | * | 0.0281 |
| Ensemble vs. Tree-based | 0.08763 | 0.01507 to 0.1602 | Yes | * | 0.0186 |
| Ensemble vs. Discriminant analysis | 0.01721 | -0.03763 to 0.07206 | No | ns | 0.534 |
| Ensemble vs. Neural network | 0.09363 | 0.02107 to 0.1662 | Yes | * | 0.0121 |
| Multiple comparison of ROC AUC scores | | | | | |
| Fisher's LSD | **Mean Diff.** | **95.00% CI of diff.** | **Significant?** | **Summary** | **Individual P Value** |
| Ensemble vs. Logistic regression | 0.09847 | 0.03259 to 0.1644 | Yes | ** | 0.0039 |
| Ensemble vs. Gaussian process | 0.2902 | 0.2243 to 0.3561 | Yes | **** | <0.0001 |
| Ensemble vs. Other linear models | 0.2152 | 0.1721 to 0.2584 | Yes | **** | <0.0001 |
| Ensemble vs. Naïve bayes | 0.03127 | -0.03461 to 0.09715 | No | ns | 0.3477 |
| Ensemble vs. Nearest neighbor | 0.2361 | 0.1702 to 0.3019 | Yes | **** | <0.0001 |
| Ensemble vs. Support vector machines | 0.1313 | 0.06540 to 0.1972 | Yes | *** | 0.0002 |
| Ensemble vs. Tree-based | 0.06816 | 0.002276 to 0.1340 | Yes | * | 0.0428 |
| Ensemble vs. Discriminant analysis | 0.01568 | -0.03412 to 0.06549 | No | ns | 0.5326 |
| Ensemble vs. Neural network | 0.1023 | 0.03643 to 0.1682 | Yes | ** | 0.0027 |
